# Predictors of treatment outcomes in adults with drug–sensitive Tuberculosis in Maharashtra, India: A retrospective study

**DOI:** 10.64898/2026.05.12.26352988

**Authors:** Raghavan Parthasarathy, Yashi Raj, Nabanita Majumder, Mithun Mitra, Sarika Mehra, Raghuram Rao, Shobini Rajan

**Author notes:** Corresponding author details: Email ID, Email ID.

## Abstract

**Background:** Tuberculosis (TB) remains the leading infectious cause of death worldwide, with India accounting for nearly one-fourth of global TB cases. Ni-kshay, the country’s digital case-based TB notification platform is rich in data pertaining to the continuum of care of TB patients. This study aims to develop a standardized analytical approach to programmatic data to identify predictors of unfavourable treatment outcomes and mortality among adult drug-sensitive TB patients at the state level for Maharashtra during 2021 and 2022.

**Methods:** Two separate analyses were undertaken comparing treatment success with: (1) unfavourable outcomes (death, treatment failure, loss to follow-up, regimen change, or not evaluated); and (2) mortality. Multivariate logistic regression was used to compute adjusted odds ratios (aOR) for key risk factors, adjusting for age, gender, and weight.

**Results:** The final cohort included 323,124 cases for unfavourable outcome analysis and 315,579 cases for mortality analysis. Increasing age, male gender, lower body weight, known HIV and diabetes comorbidities, tobacco and alcohol consumption, and "unknown" status for behavioural risks and comorbidity status were significantly associated with increased odds of both unfavourable outcomes and mortality.

**Conclusions:** This study highlights the utility of programmatic data in identifying high-risk TB patients and offers a reproducible analytic framework.

## Introduction

Tuberculosis (TB) continues to be the leading cause of mortality among infectious diseases. India has the highest TB burden globally, accounting for nearly one-fourth of the world’s TB cases. As per the Global TB Report 2025 the estimated number of TB-related deaths in India in 2024 was 305,041 (234,696–379,969), with a case fatality ratio (CFR) of 11% (8–15%)(1). This reflects a reduction of over ∼28% in terms of the TB deaths, against the interim milestone for 2025 as per the End TB strategy(2).

Considering this gap to end TB, the Government of India had prioritised TB elimination through its National Strategic Plan (NSP), which outlines the targets required to be achieved and the timelines for ending TB as a public health problem in the country(3). Central to the NSP is the National TB Elimination Programme (NTEP), formerly known as the Revised National Tuberculosis Control Programme (RNTCP), which ensures the provision of free diagnostic and treatment services nationwide. In support of effective surveillance and programmatic monitoring, the Government of India introduced the Ni-kshay platform in 2012, a digital, case-based TB notification and management system covering both public and private health sectors.

Ni-kshay, means “end (*Ni*) tuberculosis (*kshay*),” symbolises India’s commitment to defeat tuberculosis (TB). The evolution of Ni-kshay has coincided with expanded programmatic support measures, such as the introduction of the Ni-kshay Poshan Yojana (NPY), launched in 2018, which provides TB patients with INR 1000 per month to mitigate nutrition-related vulnerabilities, Pradhan Mantri TB Mukt Bharat Abhiyaan (PMTBMA, 2022) where TB patients in need are linked with community supporters willing to provide nutritional support to the affected households through Nikshay Mitras(4,5). Ni-kshay has evolved into a repository of rich, patient-level data encompassing clinical, demographic, and treatment-related variables across India’s vast and diverse populations(6).

Programmatically, treatment success is a key indicator for monitoring the effectiveness of TB treatment services and tracking progress towards the targets set under the End TB Strategy, including the reduction of TB-related mortality(7). According to the India TB Report 2024, across the country, ∼6.2% of TB patients experienced unfavourable outcomes (i.e. deaths: 3.6%, loss-to-follow-up:2.1%, treatment failure:0.5%)(8). Understanding and addressing the determinants of these outcomes is critical for meeting NSP’s vision.

This is a retrospective observational study focused on adult patients (≥15 years of age) diagnosed with drug-sensitive TB (DS-TB) and treated in Maharashtra, a high-TB burden state, during the years 2021 and 2022. The study aims to systematically analyse Ni-kshay data to identify predictors of unfavourable treatment outcomes as well as mortality using a robust, reproducible statistical framework. Through this effort, we aim to provide a replicable model including a reporting framework for Ni-kshay-based statistical analysis.

### Methodology

This was a collaborative initiative between the National Disease Modelling Consortium (NDMC), at the Indian Institute of Technology, Bombay (IIT-B)(9) and the Central TB Division (CTD), Government of India (GoI). This was a retrospective observational study utilizing routinely collected programmatic data to standardize the procedure for understanding the determinants of unfavourable treatment outcomes and mortality among adult (≥15 years) DS-TB patients from Maharashtra in 2021 and 2022.

### General setting and data source

The state of Maharashtra was purposively selected for this analysis in consultation with the Central TB Division (CTD), based on its high tuberculosis (TB) burden. The extraction methodology from Ni-kshay applied filters based on “diagnosing facility” and “notification date” for the current case study. To focus the analysis on treatment outcomes, only cases with documented treatment initiation were included. Specifically, records lacking a valid entry under the “spectrum treatment initiation date” variable, either blank or marked as “NA”, were excluded from the study cohort, as these represented individuals not initiated on treatment. Given the longer and more complex treatment timelines for drug-resistant TB (DR-TB), this study was limited to patients diagnosed with drug-sensitive TB (DS-TB). The objective was to assess determinants of treatment outcomes within a programmatic cohort for whom standard treatment timelines and outcome measures apply within a span of 12 months. It was decided to use only the notification register and comorbidity register in Ni-kshay as the primary data-source, considering the relevance for building the case-study (extracted in September, 2024). The study focused on cohorts from 2021 and 2022, as these represent completed treatment cycle cohorts with outcomes available for analysis as well as in accordance with national programmatic guidance, which recommends outcome evaluation 12 months after diagnosis. Cohorts from 2023 and 2024 were excluded, as treatment for a substantial proportion of these cases was either ongoing or not yet eligible for outcome assessment at the time of data extraction in September 2024(7).

### Specific setting and definitions

Two outcome-based comparisons were conducted (1) treatment success versus unfavourable TB treatment outcomes and (2) treatment success versus mortality. Treatment success was defined as a final outcome of "cured" or "treatment completed." Mortality referred to outcomes marked as "died."

Unfavourable outcomes included "died," "loss to follow-up," "failure," "treatment regimen changed", and "not evaluated." Recently, countries and their research programmes have begun to categorise “not evaluated” under “unfavourable outcomes”(10–12). To align with NTEP definitions(7), we would also be conducting a sensitivity analysis, excluding those individuals with the outcome of “not evaluated” from the unfavourable outcomes cohort. The counting of the number of cases was done using the variable “*instance*”.

### Risk factor/determinants

The risk-factors considered for analysis considering the completeness and correctness of the information captured were as follows: demographic variables: age (15-99 years), gender; anthropometric variable for capturing nutritional status: weight (Kg); TB related – site of disease (pulmonary or extra-pulmonary); clinical comorbidities: diabetes status, HIV status; and behavioural risk factors: tobacco consumption status and alcohol consumption status. The risk factors that were available as integers were categorized and those which were non-numerical were recoded for analysis. Height was not utilized because of the non-availability of the information in more than 1/3^rd^ of the records. To ensure completeness in the dataset, those cases where weight was not available, were excluded from analysis. In all other cases, the missing variables were categorized as “unknown” and utilized in the analysis to ensure maximum representativeness.

### Analytical procedure including data pre-processing

The comorbidity register was merged with the notification register by matching using the variable “instance”. After merging, the cases having age less than 15 years and drug-resistant TB cases (*DRTB case definition: cases with a value of “1” in the variable “RR resistance detected” or “INH resistance detected”*) were excluded. The categorical variables were described as percentages, and the Chi-square or Fisher’s exact test was used as the test of statistical significance. For categorical variables, we estimated Cramér’s V statistic to quantify the strength of association between pairs of variables (collinearity). Univariate and multivariate logistic regression analysis, using STATA version 19 (StataCorp. 2025. *Stata Statistical Software: Release 19*. College Station, TX: StataCorp LLC) was done to report the risks of various factors and reported as “odds ratio (OR)” and “adjusted odds ratio (aOR)”, respectively, with 95% confidence intervals. In the multivariate regression, all the risk factors were adjusted for age, gender and weight.

### Ethical approval

Since the study utilised the non-patient identifiable, secondary data from the programmatic database, no ethical approval was required for the study. For the overall project on TB being carried out by the NDMC and IIT-B, ethical approval was obtained from the Institutional Review Board (IRB) at IIT-B (IRB-2025-028_SM_M). Also, the analysis was conducted with post-administrative approval from the Central TB Division, NTEP, GoI. The manuscript preparation was performed in accordance with the Strengthening the Reporting of Observational Studies in Epidemiology (STROBE) statement.

## Results

The final dataset included in the analysis comprised TB cases notified in Maharashtra between 2021 and 2022. Following exclusions based on predefined criteria (see *Figure-1: Data pre-processing flow*), 323,124 cases were included in the final cohort for the analysis of unfavourable treatment outcomes, and 315,579 cases were analysed for TB-related mortality (excluding cases with outcomes other than “DIED” from the unfavourable outcomes).

**Figure 1:**
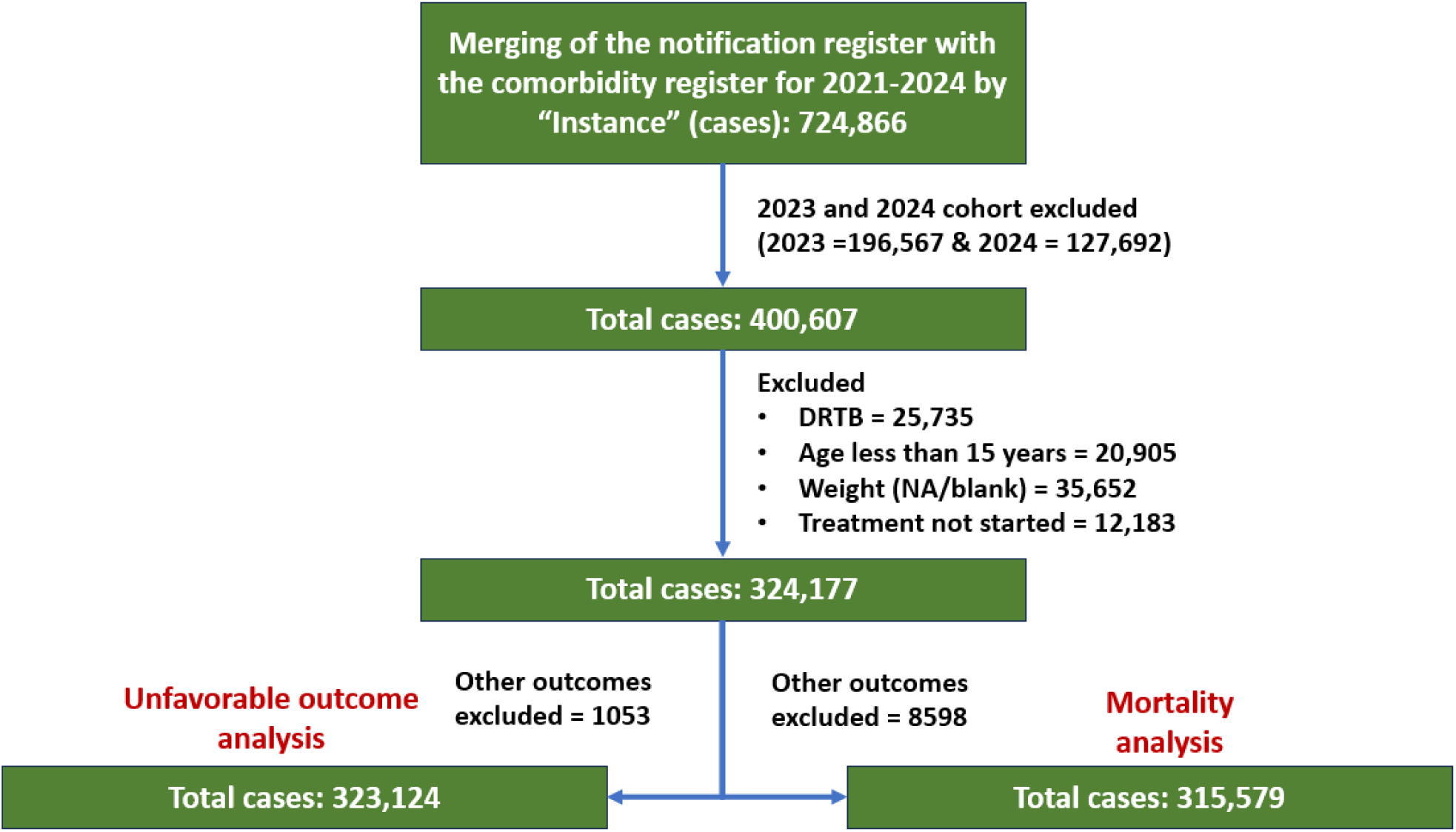
Data flow and steps in pre-processing.

### Demographic Characteristics (Table-1 & 2; Figure-2; Supporting information S5 & S6 table)

**Figure 2:**
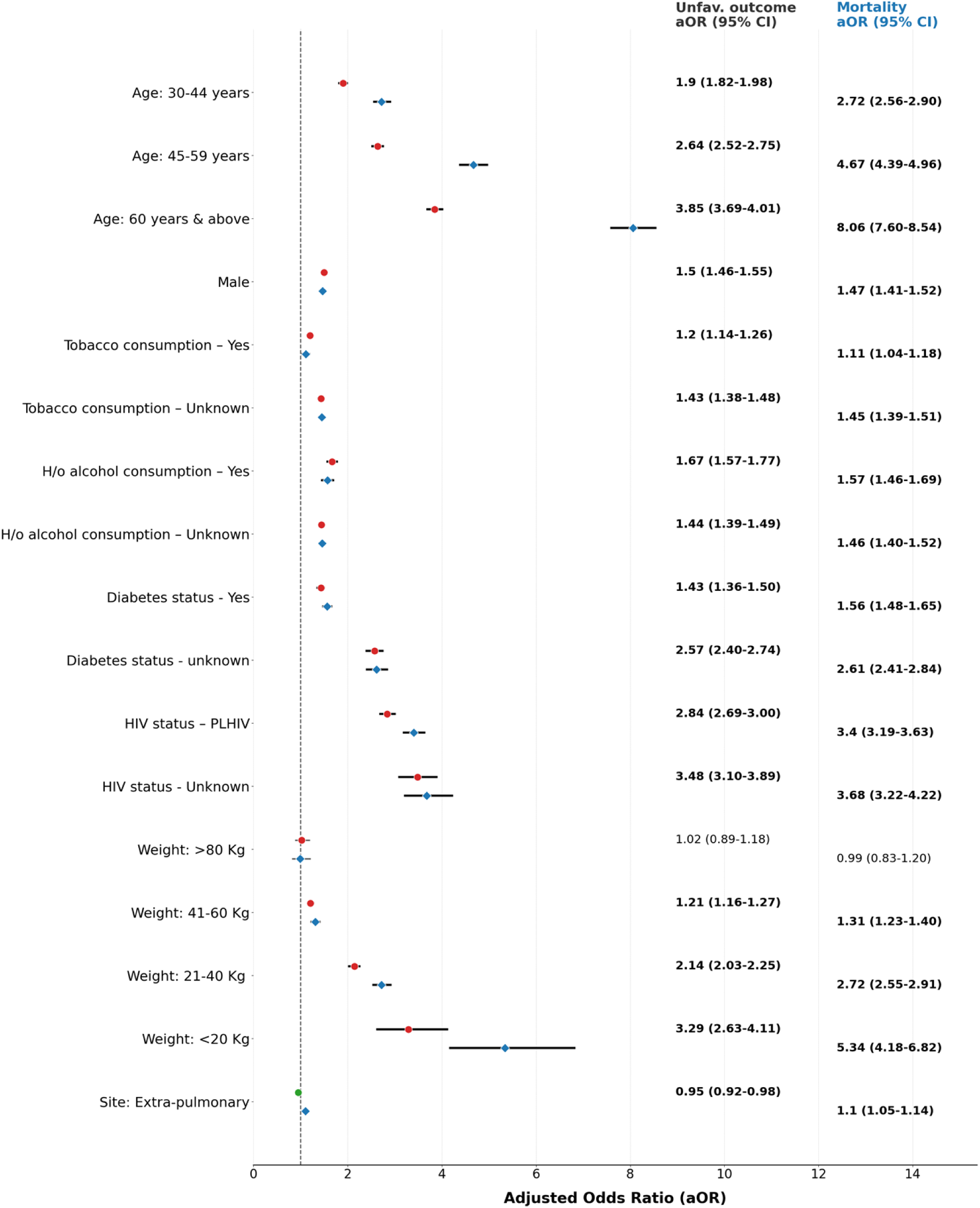
Forest plot of aOR of key risk factors against unfavourable TB treatment outcomes (Red) and TB mortality (Blue)

**Table 1:**
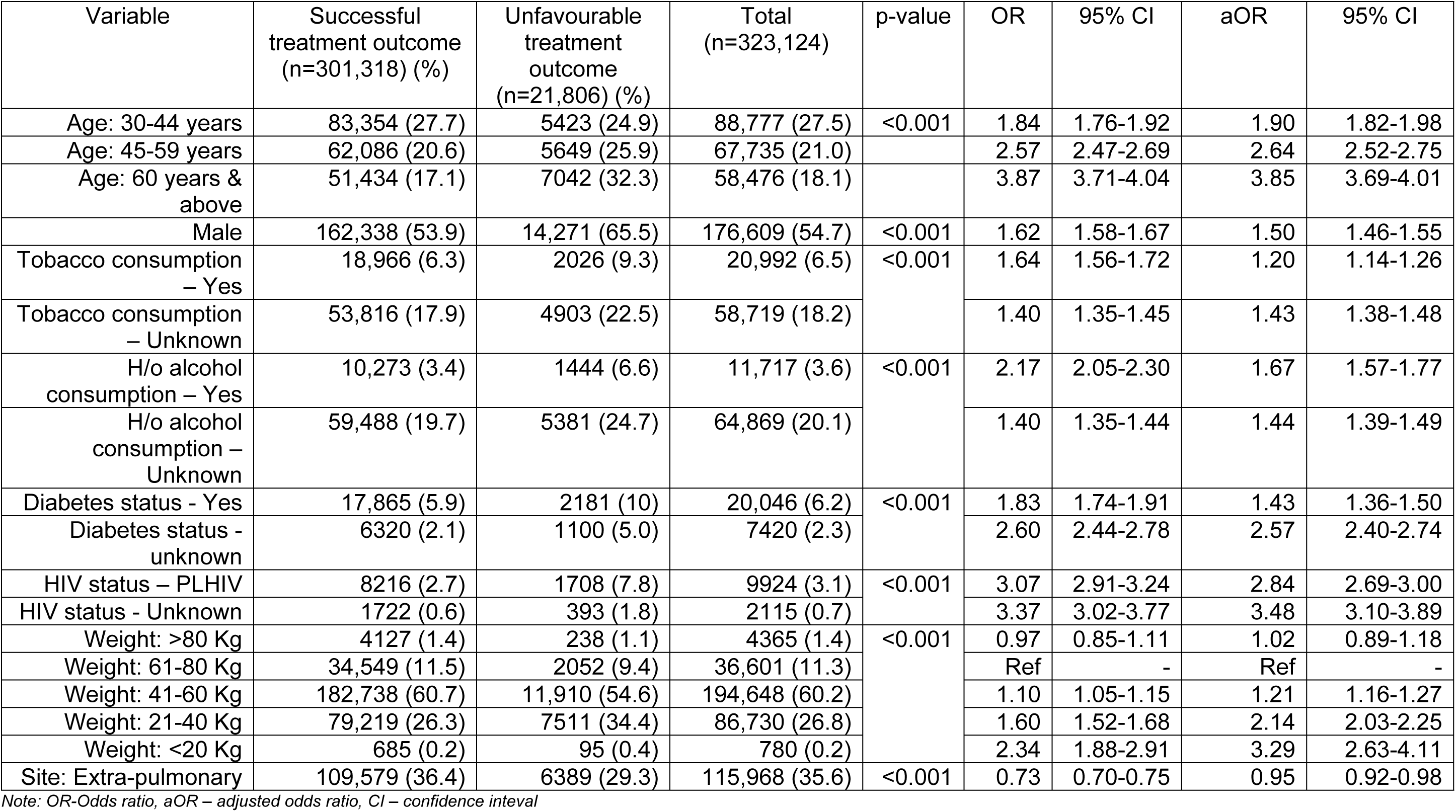
Descriptive and inferential analysis on the risk factor for unfavourable treatment outcomes from Ni-kshay for Maharashtra (2021 and 2022)

The mean (SD) age of patients in the unfavourable outcomes cohort was 48.9 (17.9) years, and 53.1 (17.4) years in the mortality cohort. The burden of both unfavourable outcomes and mortality was markedly higher among individuals aged ≥60 years, who accounted for 32.3% of all unfavourable outcomes and 40.6% of all deaths. Similarly, males were disproportionately affected, comprising 65.5% of the unfavourable outcome group and 66.2% of deaths. Multivariate logistic regression analysis indicated a consistent and significant increase in risk with advancing age. Patients aged more than 60 years had the highest adjusted odds ratios (aORs) for both outcomes: 3.85 (95% CI: 3.69–4.01) for unfavourable outcomes and 8.06 (95% CI: 7.60–8.54) for mortality. Gender-based disparities were evident; males had elevated risk (aOR: 1.50 [1.46–1.55] for unfavourable outcomes; 1.47 [1.41–1.52] for mortality), and transgender individuals demonstrated substantially higher risk (aOR: 2.52 [1.59–4.00] for unfavourable outcomes; 2.63 [1.51–4.61] for mortality).

### Behavioural risk factors (Table-1 & 2; Figure-2; Supporting information S5 & S6 tables)

Both tobacco and alcohol consumption were associated with unfavourable outcomes as well as mortality. Tobacco users accounted for 9.3% of the unfavourable outcome cohort and 9.2% of the mortality cohort. Alcohol consumption was reported in 6.6% and 6.4% of these respective cohorts. Notably, the unknown status for both tobacco (22.5% for unfavourable outcomes; 22.6% for mortality) and alcohol consumption (24.7% for both outcomes) remained high. Among categorical variables, Cramér’s V statistic indicated a strong association between tobacco consumption and alcohol consumption (0.71), reflecting a strong positive relationship whereby individuals reporting tobacco consumption were substantially more likely to also report alcohol consumption. However, we retained both variables in our models, given their potential for independent biological effects on TB pathogenesis and outcomes. Regression analyses showed a higher risk associated with alcohol consumption (aOR: 1.67 [1.57–1.77] for unfavourable outcomes; 1.57 [1.46–1.69] for mortality than with tobacco consumption. While known tobacco consumption was linked to elevated risk, those with unknown tobacco status had slightly higher aORs (1.43 [1.38–1.48] for unfavourable outcomes; 1.45 [1.39–1.51] for mortality).

### Comorbidities (Table-1 & 2; Figure-2; Supporting information S5 & S6 tables)

Among the cases, diabetes was reported in 10% of those with unfavourable outcomes and 11.6% of those who died. PLHIV constituted 7.8% and 9% of the respective groups. Both diabetes and HIV status were significant predictors of unfavourable outcomes, even after adjusting for demographic variables and weight. Individuals with unknown HIV or diabetes status also exhibited increased risk. Lower body weight emerged as a significant predictor of both unfavourable treatment outcomes and mortality among TB patients. Individuals in the 41–60 kg weight category constituted the majority of those with unfavourable outcomes, representing 54.6% of the unfavourable outcome group and 52.6% of those who died. A clear dose-response relationship was observed, with the adjusted odds ratio (aOR) for adverse outcomes increasing as body weight declined. Patients weighing less than 40 kg were found to be at the highest risk, indicating a strong vulnerability. An analysis stratified by age and weight revealed distinct shifts in the demographic and clinical profile across outcome categories (see Supporting information S4 table). The cohort with successful treatment outcomes was predominantly composed of younger to middle-aged individuals (15–59 years) within the 41–60 kg weight range. However, in the group with unfavourable outcomes, there was a marked increase in the proportion of older adults (≥60 years), who accounted for 32.3% of this subgroup, along with a higher prevalence of individuals in the severely low weight range (21–40 kg). This pattern was even more pronounced in the mortality cohort. Patients aged 60 years and above contributed to over 40% of all recorded deaths, while more than 90% of the deaths occurred among those weighing less than 60 kg. These findings highlight a synergistic and compounding risk posed by advanced age and low body weight, underscoring their critical role in the transition from treatment success to unfavourable outcomes.

### TB disease site and outcome relationship (Table-1 & 2; Figure-2; Supporting information S5 & S6 tables)

Cases with extra-pulmonary TB (EPTB) demonstrated lower proportions of both unfavourable outcomes and mortality. Interestingly, multivariate analysis revealed a divergent trend where EPTB was protective against unfavourable outcomes but conferred an increased risk of mortality. The temporal trends across 2021–2022 remained largely stable (Supporting information S1, S2 and S3 tables), and the comprehensive versions of Table-1 and Table 2 are presented in supporting information S5 & S6 tables.

**Table 2:**
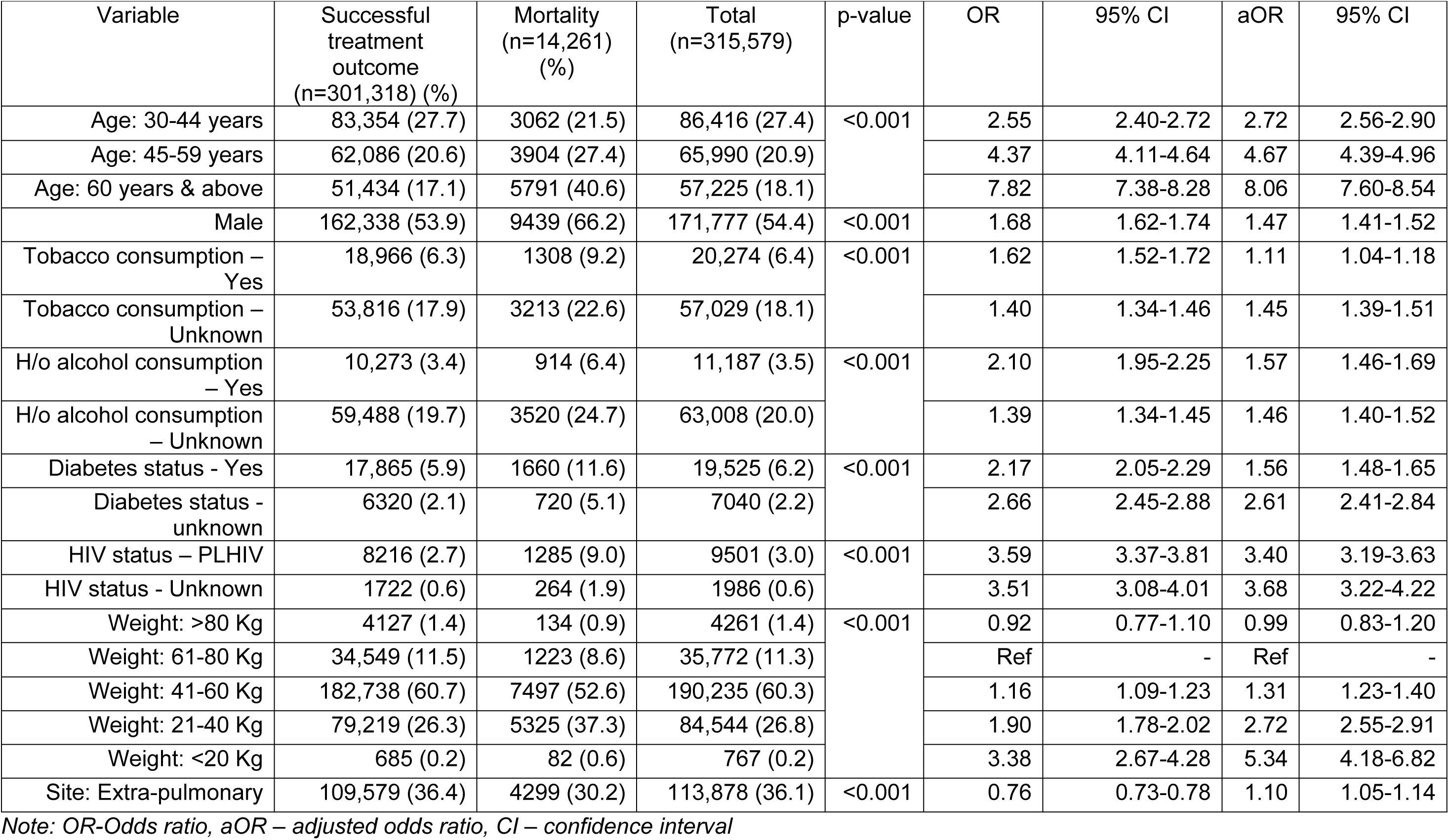
Descriptive and inferential analysis on the risk factor for TB mortality from Ni-kshay for Maharashtra (2021 and 2022)

### Sensitivity analysis

Four sensitivity analyses were carried out to ensure the robustness of risk factor associations for unfavourable treatment outcomes as well as mortality. First, we created a composite substance use variable combining tobacco and alcohol consumption to address collinearity, which was observed yet retained considering the different biological pathways for tobacco and alcohol consumption. Second, we compared baseline characteristics between patients with complete versus missing weight. Third, we performed multiple imputation using predictive mean matching with 5 nearest neighbours (k=5) to assess the impact of missing weight data on association (aOR) estimates. We generated 10 imputed datasets using age group, gender, diabetes status, HIV status, substance consumption, and site of TB disease as predictors). Fourth, we excluded all patients classified as ’not evaluated’ from the unfavourable outcome category and repeated the overall multivariate analysis.

The composite substance use variable eliminated collinearity, with all pairwise correlations below 0.32 (*Supporting information* S7 table). Missing data versus complete case comparison revealed that ∼6% of patients lacked weight information with similar distribution of variables between the two groups (*Supporting information S5 & S6 tables* S8a, S8b tables). Multiple imputation using predictive mean matching produced consistent results across all 10 imputations (both in the unfavourable outcome and mortality comparison), with no impossible values generated. In multivariable models, for unfavourable outcomes and mortality the aOR for factors showed consistent associations across methods, indicating that missing weight data did not substantially bias findings (*Supporting information* S9 table) as compared to the primary analysis (Table: 1 & 2). Finally, upon exclusion of all patients classified as ’not evaluated’ from the unfavourable outcome category and repeating the overall analysis, the aORs for all risk factors remained virtually unchanged (not shown), confirming that this category (n=513) in the current unfavourable outcomes cohort did not affect the observed associations.

## Discussion

This study represents one of the first large-scale attempts to apply a standardized analytical protocol to programmatic data for the retrospective analysis of unfavourable treatment outcomes and mortality among adult drug-sensitive TB (DS-TB) patients. By using routine programmatic data from Maharashtra, a high-burden state(8), across two calendar years (2021 and 2022), we demonstrated the value of programmatic data both as a surveillance and decision-support tool when methodologically harnessed. Our analysis affirmed known risk factors(13–18), both from India and worldwide: age, sex, HIV, diabetes and current tobacco and alcohol consumption. Males, older adults (especially above 60), and patients with co-morbidities like diabetes or HIV had higher odds of both unfavourable outcomes and mortality after adjustment. Also, for Maharashtra, we observed that the risk due to HIV was lower than observed across another study in Central India(19), possibly because of the higher coverage of TB-HIV screening and antiretroviral treatment initiation in the region(20).

Notably, the “unknown” status for tobacco, alcohol, diabetes, or HIV(17) emerged as a significant risk category in itself. This finding reinforces the critical importance of high-quality data entry and the need for frontline worker training in holistic risk assessment(21). From a health systems perspective, the “unknown” category in Ni-kshay could arise from one of several scenarios: (a) the screening test was not offered or not performed at the diagnosing facility; (b) the test was performed but the result was not entered into the Ni-kshay portal, reflecting a data entry gap rather than a clinical gap; (c) the patient was diagnosed and notified by the provider, where integration with NTEP comorbidity screening protocols has historically been weaker; or (d) the patient declined testing due to stigma or logistical barriers. Each of these scenarios represents a distinct point of programmatic challenge that need to be addressed through targeted interventions. This finding aligns with the observations from the assessment of Ni-kshay(6). However, considering the COVID-19 pandemic and the various constraints within which programmatic services were delivered during 2021 and 2022, including disruptions to routine screening, the completeness of information was understandably lower than what would be expected and should be interpreted with this operational context in mind(8).

One of the more nuanced findings was the role of weight as a predictive factor. Patients with low baseline body weight had significantly higher risks of unfavourable outcomes, supporting earlier studies highlighting malnutrition as a core determinant of TB mortality(19,22–24). Approximately a quarter of the adult DSTB patients in Maharashtra had a pre-treatment body weight of less than 40 kg, comparable to other studies(25), making the findings generalizable to the overall population. Given the known association between undernutrition and TB outcomes in India, there is a compelling need to integrate and monitor regular anthropometric assessments and the findings have practical relevance in the context of the recently released National Guidance on Differentiated TB Care, which recommends risk stratification and severity assessment of all notified patients at diagnosis(26). Substance use (tobacco or alcohol consumption) that was found to be associated with unfavourable outcomes could be a potential triaging variable. Patients identified with substance use could be flagged within Ni-kshay for enhanced adherence support, treatment monitoring, more frequent clinical review, and referral linkage to de-addiction services where available. This approach aligns with the Risk and Needs Assessment (RNA) tool piloted under the THALI project in Karnataka and Telangana, which identified substance use as a key predictor of poor outcomes(27).

Another observation from the study was the divergent relationship between EPTB and the two outcome measures. EPTB was associated with lower odds of unfavourable outcomes overall (aOR: 0.95, 95% CI: 0.92–0.98) but with slightly higher odds of mortality specifically (aOR: 1.10, 95% CI: 1.05–1.14). The slightly elevated mortality risk among EPTB patients likely reflects a combination of diagnostic delays inherent to the non-specific presentation of extrapulmonary disease, the involvement of high-fatality anatomical sites and differential age-groups getting. This finding is consistent with evidence from a large Chinese cohort in which age above 60 years increased the hazard of mortality by over 11-fold among patients with concurrent pulmonary and extrapulmonary disease, and by nearly 24-fold among those with exclusively EPTB(28). Taken together, these observations support the case for including EPTB, particularly in older adults, as a criterion for early comprehensive clinical assessment and triaging within the differentiated care framework.

Despite its strengths, this study faced certain methodological challenges that have scope for improvement in future research. Future statistical modelling should expand beyond the two registers utilized here, as the programmatic database contains a wealth of additional patient-centric information. While the exclusion of those not initiated on treatment and those with missing baseline weights might have underestimated the risks, the sensitivity analyses performed through multiple imputation confirms the robustness of our primary findings. Missing data patterns (baseline body weight), while non-random, did not correlate strongly with outcomes, and multiple imputation yielded nearly consistent effect estimates comparable to the complete-case analysis. The consistency across analytical approaches strengthens the primary findings. Predictive mean matching, which ensures that imputed values match the observed data distribution, produced biologically plausible results without generating impossible values.

Finally, the successful merging of notification and comorbidity registers using the “instance” variable in alignment with the latest programmatic norms and reporting framework could be a replicable method for future analyses. Majority of the studies that have utilized programmatic data in past have provided less clarity and utilized varying methodologies on which identifier was the identifier used for counting cases and subsequent analysis(21,29–40). Hence, we also propose a standardized reporting framework (Table-3) that future researchers and program managers can adopt when conducting inferential analyses using TB-related programmatic data from Ni-kshay.

**Table 3:**
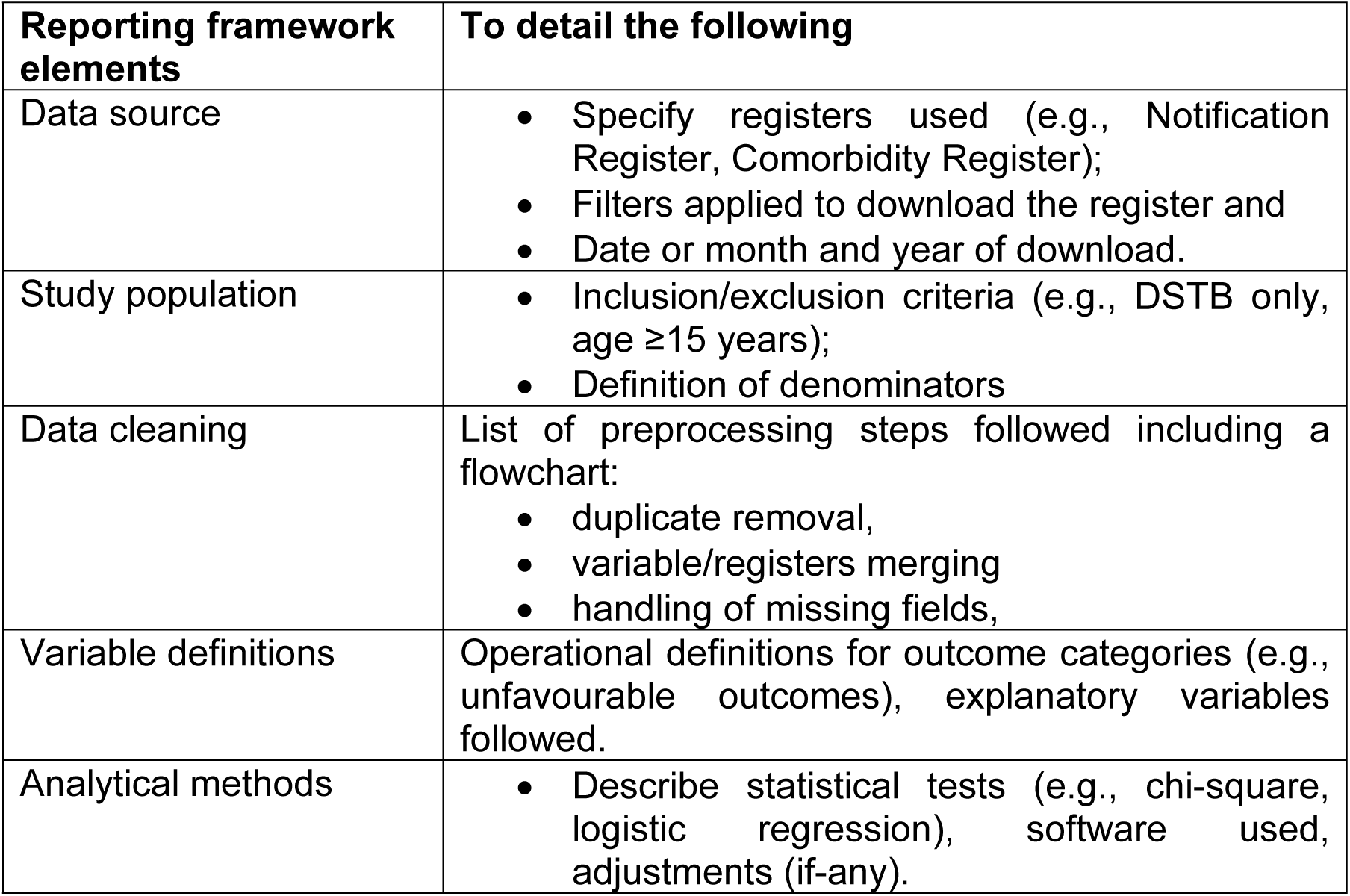
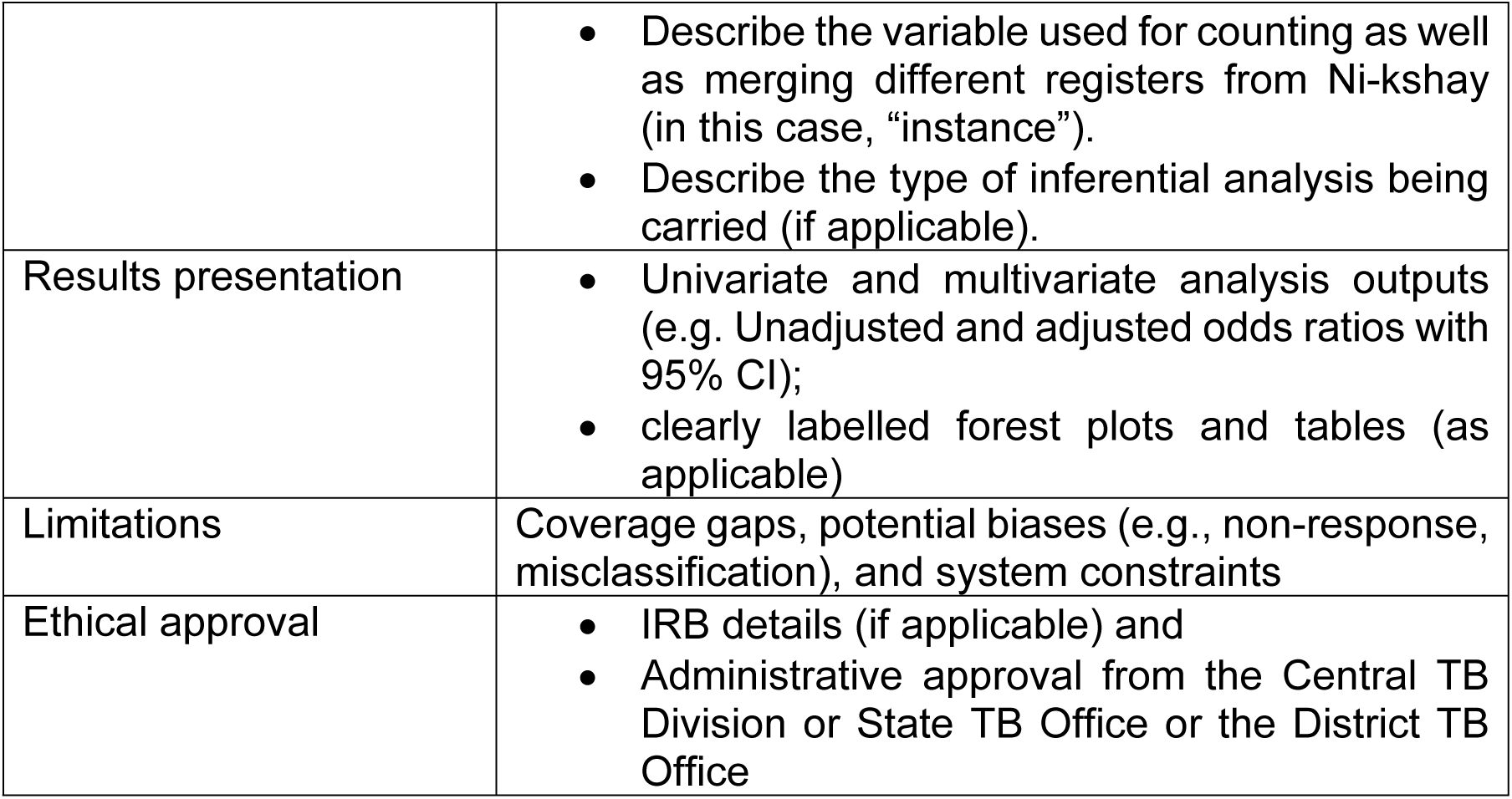
Standardized reporting framework for studies utilizing Ni-kshay-based data.

| Reporting framework elements | To detail the following |
| --- | --- |
| Data source | Specify registers used (e.g., Notification Register, Comorbidity Register);<br>Filters applied to download the register and<br>Date or month and year of download. |
| Study population | Inclusion/exclusion criteria (e.g., DSTB only, age $\geq 15$ years);<br>Definition of denominators |
| Data cleaning | List of preprocessing steps followed including a flowchart:<br>duplicate removal,<br>variable/registers merging<br>handling of missing fields, |
| Variable definitions | Operational definitions for outcome categories (e.g., unfavourable outcomes), explanatory variables followed. |
| Analytical methods | Describe statistical tests (e.g., chi-square, logistic regression), software used, adjustments (if-any). |
|  | • Describe the variable used for counting as well as merging different registers from Ni-kshay (in this case, “instance”).<br>• Describe the type of inferential analysis being carried (if applicable). |
| Results presentation | • Univariate and multivariate analysis outputs (e.g. Unadjusted and adjusted odds ratios with 95% CI);<br>• clearly labelled forest plots and tables (as applicable) |
| Limitations | Coverage gaps, potential biases (e.g., non-response, misclassification), and system constraints |
| Ethical approval | • IRB details (if applicable) and<br>• Administrative approval from the Central TB Division or State TB Office or the District TB Office |

## Conclusion

This study offers a robust, replicable analytical framework grounded in demographic, clinical, and behavioural indicators for understanding the predictors of treatment outcomes using the programmatic data adhering to the updated norms, guidelines and definitions. Future work should expand this analysis to other Indian states and incorporate more recent data from 2023 and 2024. Furthermore, investigating the impact of additional interventions available from the programmatic database, such as direct benefit transfer, treatment support systems and patient adherence levels using advanced statistical methods, such as Poisson, Cox regression, and predictive modelling, can enable real-time risk stratification and better inform evidence-based public health interventions, as well as monitor the impact of the newer strategies on reducing the unfavourable outcomes and mortality.

## Data Availability

The request for data with valid justification will be submitted to the programme division for approval and the same anonymized dataset will be made available on a case-to-case basis post-approval. Also, to note, the tables in the manuscript and the supporting information document has been made exhaustive to enable any additional analysis and inferences that may be of interest to the readers.

## Acknowledgements

RP, YR, NM, SM, MM and RR and SR were involved in conceptualising the study. Data analysis was carried out by RP, YR and NM, with the initial draft prepared by RP. YR, SM, MM, RR and SR provided critical review and valuable feedback on the manuscript. We extend our sincere thanks to the faculty and staff of the NDMC, IIT Bombay, for their support in conducting this study. We also acknowledge the support provided by Ms Krittika Bhattacharyya, from NDMC, for supporting in the data preparation. We also acknowledge the guidance and inputs provided by Dr Urvashi B Singh, Professor, In-Charge, Tuberculosis Division, Department of Microbiology, All India Institute of Medical Sciences, New Delhi. Funding for this study was provided by the Gates Foundation (Grant No. INV-044445). Dr. Raghavan Parthasarathy received funding support for this study from the Gates Foundation (Grant No. INV-065264).

## Disclaimer

*The work/opinion is based on research findings by the authors and not the opinion of the government*.

## Supporting information

S1 Table: Descriptive analysis of the adult, DSTB cohort for 2021 and 2022 with successful and unfavourable treatment outcomes

S2 Table: Descriptive analysis of the adult, DSTB cohort for 2021 and 2022 with successful treatment outcome and mortality

S3 Table: Adjusted* odds ratios (aOR) of various risk factors against unfavourable treatment outcomes and mortality for 2021 and 2022

S4 Table: Age and weight-wise categorisation of different outcomes (successful outcomes, unfavourable outcomes and mortality) among cases across the 2 years combined

S5 Table: Descriptive and inferential analysis on the risk factor for unfavourable treatment outcomes from Ni-kshay for Maharashtra (2021 and 2022)

S6 Table: Descriptive and inferential analysis on the risk factor for TB mortality from Ni-kshay for Maharashtra (2021 and 2022)

S7 Table: Pairwise Cramér’s V correlations after creating a composite substance use variable for addressing collinearity of exposure variables for the two outcomes

S8a Table: Description of baseline characteristics between complete and missing weight data: Unfavourable outcomes dataset (N = 343,479)

S8b Table: Description of baseline characteristics between complete and missing weight data: Mortality dataset (N = 335,334)

## Notes

### Competing Interest Statement

The authors have declared no competing interest.

### Author Declarations

Since the study utilised the non-patient identifiable, secondary data from the programmatic database, no ethical approval was required for the study. For the overall project on TB being carried out by the NDMC and IIT-B, ethical approval was obtained from the Institutional Review Board (IRB) at IIT-B (IRB-2025-028_SM_M). Also, the analysis was conducted with post-administrative approval from the Central TB Division, NTEP, GoI.

